# Association between obesity, brain atrophy and accelerated brain aging and their genetic mechanisms

**DOI:** 10.1101/2022.12.30.22284052

**Authors:** Jujiao Kang, Tianye Jia, Zeqiang Linli, Yuzhu Li, Wei Cheng, Shuixia Guo, Jianfeng Feng

## Abstract

**Objective:** To investigate the causal relationship and the underlying biological mechanisms between body mass index (BMI) and grey matter volume (GMV).

**Methods:** We applied Mendelian randomization analyses utilizing 33,6514 individuals from the UK Biobank cohort to establish the causal relationship between BMI and GMV. We also quantified obesity-related accelerated brain aging using an XGBoost prediction paradigm. Then, mediation analyses were performed to test the association between BMI, brain atrophy, brain aging and cognitive function. Finally, the gene expression data from the Allen Human Brain Atlas were used to identify genes contributing to the BMI-GMV association.

**Results:** A causal effect of increased BMI on decreased GMV was established using multiple Mendelian randomization methods. The brain age prediction paradigm achieved appreciable performance in both training (R = 0.725, mean-absolute-error (MAE) = 4.130) and test data (R= 0.71, MAE = 4.239). On average, overweight and obese individuals exhibited significantly accelerated brain aging by +0.59 years and +1.7 years, respectively. Further, the accelerated brain age and total GMV mediated 18% of the association of higher BMI with poorer cognitive function. BMI-associated lower GMVs were related to the over-expression of gene TRIM27 and other genes involved in the autophagy biological process.

**Conclusion:** Obesity led to GMV decline and accelerated brain aging. Genes including TRIM27 and biological process pathways involved in autophagy may contribute to the BMI-GMV association.

## Introduction

Obesity is a global health problem that exacerbates organ aging and is associated with many health problems and diseases [1], including cognitive decline [2] and dementia [3,4]. Recent observational studies have reported smaller whole brain volumes in healthy but obese individuals [5,6]. However, few studies have investigated the causality and mechanisms of the association between obesity and brain structure.

Mendelian randomization (MR) is a powerful causal inference method that utilizes genetic variations as natural experiments by assigning individuals to different groups based on the segregation of alleles, hence forming a randomized experiment [7]. A properly conducted MR could minimize issues of residual confounding and reverse causation and have previously been applied to examine the causal effects of obesity [8,9]. However, horizontal pleiotropy, i.e. a single genetic variation, or a group of linked genetic variants, could underlie multiple phenotypes simultaneously, i.e. affecting the outcome not necessarily through its impact on the exposure, thus limiting the interpretation of MR results. Here, we proposed an improved MR approach to minimize the pleiotropy influence by introducing a non-pleiotropic polygenic risk score as an instrumental variable [10], which was then employed to investigate mutual causalities between obesity and brain structure.

Although many studies have explored the relationship between obesity and brain structure, few studies have quantified the influence of obesity on brain aging. Also, previous studies have attributed age-related brain atrophy in older adults [11,12] to different modifiable factors including obesity [13,14]. Therefore, unravelling the impact of obesity on pathological trajectories of brain aging has important implications for possible treatments of age-related diseases. It has recently been proposed to measure accelerated brain aging using the brain age gap (BAG), i.e. the difference between the chronological age since the date of birth and the brain age estimated from neuroimaging data [15-18], in multiple diseases such as Alzheimer’s and Parkinson’s [18,19].

In this study, we used data from UK Biobank [20], one of the largest neuroimaging databases, to investigate the relationship between obesity, brain structures and brain aging. Particularly, we assessed the causal relationship between body mass index (BMI) and brain structure by Mendelian randomization. We further explored the neurobiological mechanisms underlying the whole-brain causal influence of BMI using the brain-wide gene expression data from Allen Institute for Brain Science [21]. Finally, we quantified obesity-induced accelerated brain aging using an advanced machine learning paradigm.

## Materials and Methods

### Study participants

Study participants were selected from the UK Biobank study [20], where we restricted the samples to white British to enable valid Mendelian randomization and excluded individuals diagnosed with Alzheimer’s or dementia indicated by codes G30/F00 in ICD-10. Eventually, 18416 individuals from the first wave of the brain magnetic resonance imaging (MRI) data were included as the discovery sample, and the newly-released 15709 individuals from the second wave of the MRI data were used as an independent validation sample. Table 1 summarizes relevant demographic information. The protocols of behavioural and neuroimaging data collection are publicly available [20,22]. All participants have provided written informed consent to the UK Biobank. The UK Biobank study received ethical approval from the NHS National Research Ethics Service North West (reference number: 16/NW/0274).

### Association analysis

We used a linear model to examine the association of BMI with grey matter volume (GMV), with BMI as the predictor, and covariates including age, gender, dummy coded imaging sites, and total intracranial volume (TIV). Similarly, associations between BMI, GMV and cognitive functions (see Supplementary Materials for details) were tested. Finally, we also investigated the associations of the BAG with cognitive functions, with the BAG as the predictor, and with adjustments of age, gender, dummy-coded imaging sites, and total intracranial volume (TIV).

### Mendelian randomization (MR)

Using the availability of neuroimaging data as a random stratification of UK Biobank data, we conducted a GWAS of BMI in the large discovery sample without neuroimaging data and used MR to infer the causal relationship of BMI with GMV in the resting sample with neuroimaging data. Similarly, we recruited complete neuroimaging data to conduct a GWAS of TGMV and checked for the causal inference of TGMV on BMI in the data without neuroimaging.

We first employed the TwoSampleMR R package (https://github.com/MRCIEU/TwoSampleMR) to perform a two-sample Mendelian randomization to estimate the causal influence of BMI on GMV, and vice versa. In particular, SNPs with P value < 5E−08 and r^2^ < 0.001 in a 1000-kb window size were identified as genetic instruments. Then, the effect of each SNP was combined using the inverse-variance weighted meta-analysis, and the result represents the slope of a weighted regression of SNP-outcome effects on SNP-exposure effects, where the intercept is constrained to zero [23]. The weighted median meta-analysis is consistent even when up to 50% of the genetic instruments are invalid [24]. To identify potential horizontal pleiotropy, we used several methods, including the MR-Egger [25] and MR-PRSSEO [26].

We also employed the MR method based on non-pleiotropy PRS (MR-PRS method). Following the procedure from a previous study [10], we conducted a new GWAS of TGMV in the first wave of neuroimaging data (N = 14798) and calculated the PRSs of BMI and TGMV (using the P-value threshold of 0.05) in the independent second wave of neuroimaging data (N = 12867). Specially, we obtained the estimator of SNP effect based on the following regression models:

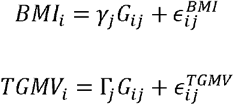

where *G*_*ij*_ = [0,1,2], *i* = 1, …, *j* = 1, …, *M* denotes the genotype of the *j*th variant from the *i*th individual, *γ*_*j*_ denotes the effect of the *j*th SNP on BMI and Γ_*j*_ denotes the effect of the *j*th SNP on TGMV. With the SNP effects estimated using individuals from the first wave of neuroimaging data., we then calculated the PRS for BMI and TGMV with a P-value threshold (PT) of 0.05 based on the following equations in the independent second wave of neuroimaging sample:

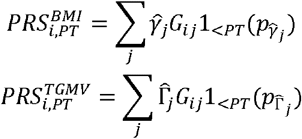

Further, we obtained the valid-PRSs by gradually removing potential pleiotropic SNPs, which cross-associated with both phenotypes, based on a predefined set of removing P-value thresholds (RT) on the following equation.

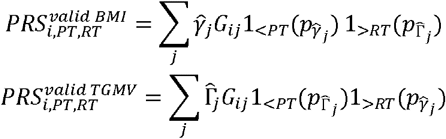

The predefined set of RT ranges from 0.05 to 0.50 with a step of 0.05. We then investigated whether the valid-PRSs were qualified instrumental variables by testing their association with the corresponding main phenotypes. Finally, the causal effect of BMI on TGMV was tested by the associations of valid-PRS_BMI_ with TGMV, and the associations of valid-PRS_TGMV_ with BMI. To examine the reproducibility of the above procedure, we also used the PRS averaging across all 10 PT (from 0.05 to 0.5, step by 0.05) or the best fit PRS as a substitute for PT = 0.05.

### GMV-based brain age prediction framework

We built a brain age predicting model based on individuals with normal body weight using the extreme gradient boosting (XGBoost) [27] predictors, i.e. a collection of gradient-boosted trees designed for speed and performance, which had performed well in recent brain age studies. We used a nested five-fold cross-validation (CV) framework, with the inner five-fold CV loop to determine the optimal parameter set and the outer five-fold CV loop to estimate the generalisability of the model. The tuning parameters include a maximum depth *M* ∈ [2,3,4,5,6,7,8,9,10], number of estimators *N* ∈ [50,100,150,200,250], the learning rate *η* ∈ [0.01,0.05,0.1,0.2,0.5]. The nested five-fold CV loop produces five XGBoost predictors with optimized parameters at each iteration, and all the training data have a single predicted age (i.e. the Brain age) based on the corresponding optimized predictor. We then used the five XGBoost predictors to predict the brain age in individuals with overweight or obesity and averaged the five predicted values as the final brain age, and the BAG was calculated as the difference between the predicted age and the chronological age (brain age - true age). To examine the prediction accuracy, we calculated the Pearson correlation coefficient (R), mean absolute error (MAE), root mean square error (RMSE) in the normal weight group and the overweight or obesity group, respectively.

### Brain age correction

As has been consistently reported[16,28], the brain-age prediction could involve certain biases, i.e. brain age is overestimated in younger individuals whereas underestimated in older individuals. and prediction performance tends to be better in individuals with age closer to the mean age of the training dataset) due to issues including regression dilution and non-Gaussian age distribution. To avoid spurious correlations between the BAG and age-related variables, we applied a regression model that included linear and quadratic effects of the chronological age to remove the effect of chronological age on the brain age as suggested by previous studies [16,29,30]. The correcting model is as the formula: *BrianAge* = *β*_0_ + *β*_1_*Age* + *β*_2_*Age*^2^ + *ε*. The corrected brain age gap is 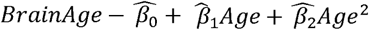 The corrected brain age is the chronological age plus the corrected brain age gap.

### Mediation analysis

Three mediation models were conducted in the current study. First, the serial mediation model was conducted to investigate whether the association between BMI and cognitive function was mediated by GMV and BAG, adjusted for age, gender, scanning sites and TIV. Further, two other mediation analyses were performed to assess whether the association between BMI and cognitive function could be mediated only by GMV or BAG, using the same covariates in the first model.

### Transcriptional correlation analysis

We examined the similarity among the BMI-linked structural association map and the gene expression map using the Pearson correlation. To ensure that the potential oversampling of brain regions will not inflate the false positive rate, we employed a *K* = 100000 permutation process to evaluate the significance level of the observed Pearson correlation coefficient *R*_*observed*_. Specifically, we shuffled the individual IDs of the GMV data at each iteration and re-calculated the Pearson correlation 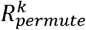 of the regenerated structural association map and the gene expression map to generate a null distribution of Pearson correlations. The corresponding p-values were hence calculated as the chance of randomly getting a correlation higher than the observation in terms of their absolute values based on the established null distribution, i.e. 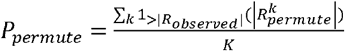.

### Analyses overview

**Figure 1**. outlines the analysis pipeline of the present study. The structural MRI was preprocessed with the standard VBM pipeline to extract GMV features using the AAL3 brain atlas. The genotyping information was obtained from whole blood. Then, several different Mendelian randomization methods were used to infer the causal association between BMI and TGMV. Meanwhile, an XGBoost brain-age prediction model was trained using a nested 5-fold CV in the normal-weight group and tested in overweight or obese individuals. The obesity-related brain aging was characterized using the BAG, which was calculated as the difference between predicted age and true age. Further, a series of association and mediation analyses were conducted among BMI, TGMV, BAG, and cognitive function. Finally, using whole-brain gene expression data, similarity analyses between GMV-association maps and whole-brain gene expression maps were performed to identify genes and biological pathways that may be related to the BMI-GMV link.

**Figure 1.**
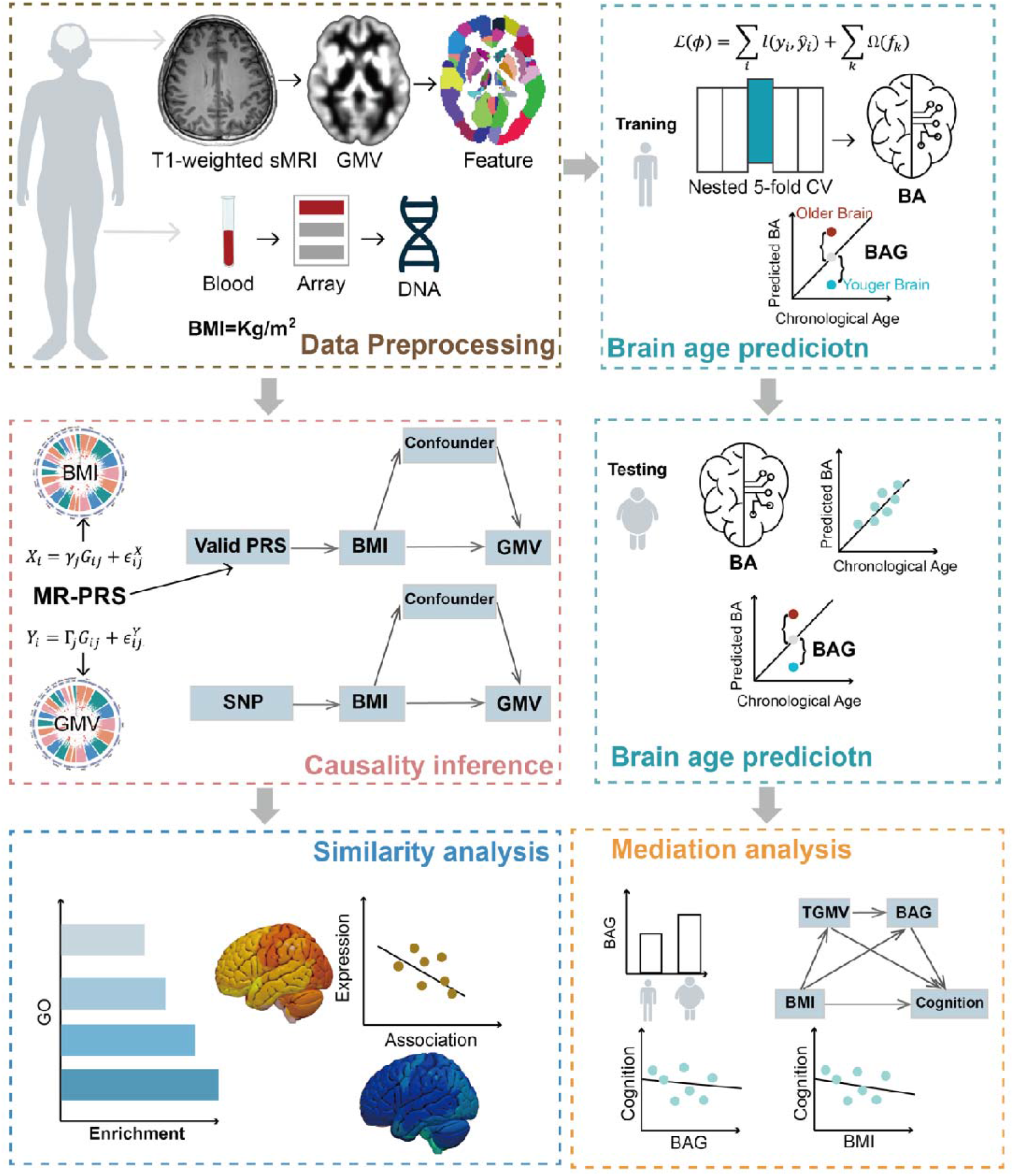
Analyses overview. The structural MRI were preprocessed with the standard VBM pipeline to extract GMV feature using the AAL3 brain atlas. The genotyping information was obtained from the whole blood. Then, several mendelian randomization methods were used to infer the causality association between BMI and TGMV. Meanwhile, a XGBoost brain-age prediction model was trained using a nested 5-fold CV method in normal-weight group and tested in overweight or obese individuals. The obesity-related brain aging was characterized using the brain age gap (BAG), which was calculated as the difference between predicted age and true age. Further, a serial of association analyses and mediation analyses between BMI, TGMV, BAG, and cognitive function were conducted. Finally, using whole-brain gene expression data, similarity analyses between GMV-association maps and whole-brain gene expression maps were performed to identify genes and biological pathways that may be related to BMI-GMV link.

## Results

### Causal effects of higher BMI on lower grey matter volume

We first utilized several established Mendelian randomization methods to investigate the potential causality between BMI and TGMV. Using the IVW meta-analysis [23] of MR based on SNP genetic instruments (see Materials and methods for details), we found a causal role of higher BMI over lower TGMV (**Figure 2A**, *β* = -1.53, se = 0.25, *P* = 8.20E-10). The same conclusion was obtained using the weighted median analysis [24] (**Figure 2A**, *β* = -1.74, se = 0.36, *P* = 1.32E-06) and the MR–Egger regression [25] (**Figure 2A**, *β* = -1.86, se = 0.71, *P* = 9.39E-03). Using the MR-PRESSO outlier-corrected test [26], the results are still significant (β = -1.49, sd = 0.23, P = 7.22E-10).

**Figure 2.**
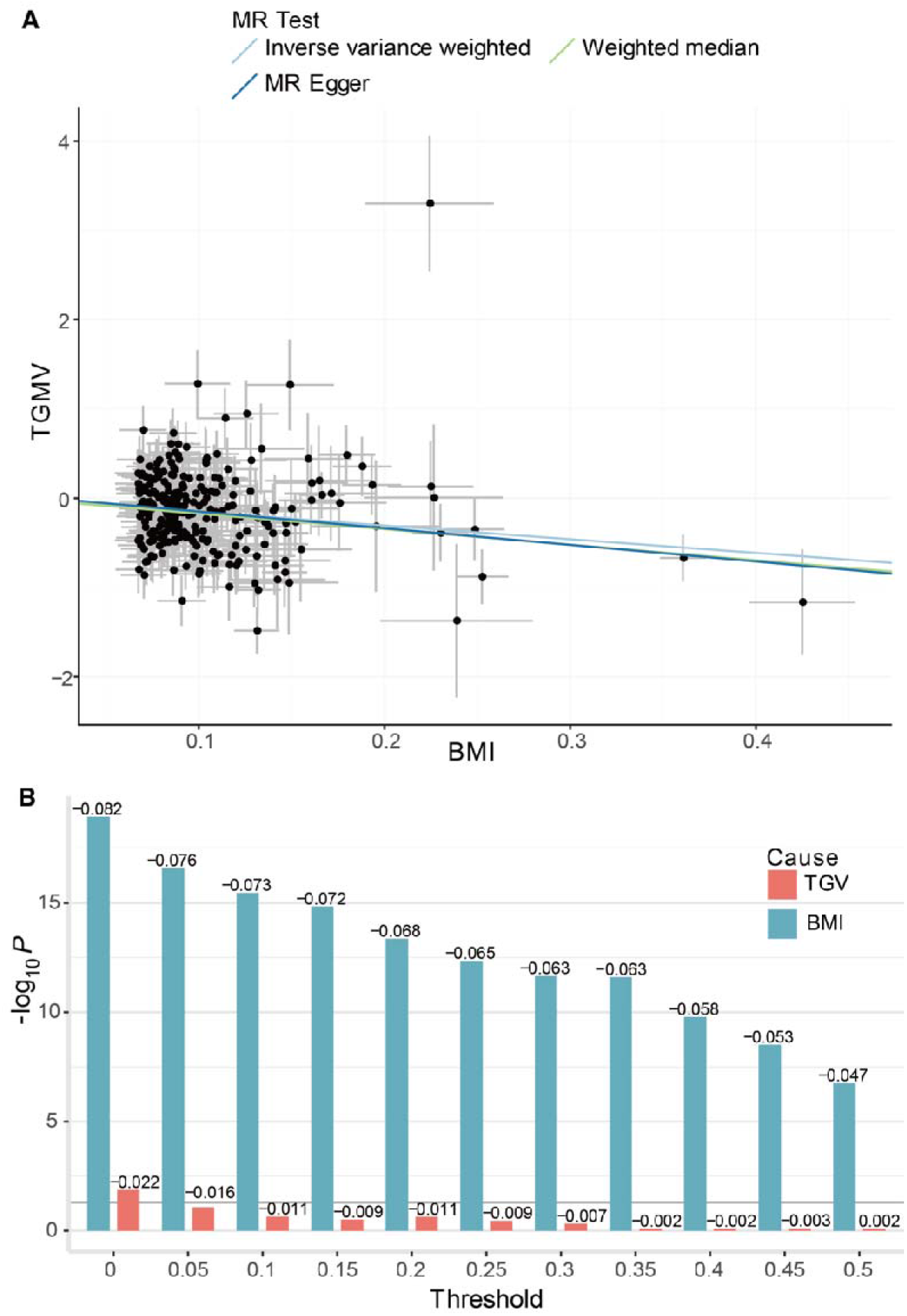
Results of Mendelian randomization analyses. **A**. Each point represents the effect estimate of each SNP and its standard error. The horizontal axis represents the effect estimates of SNP on BMI. The vertical axis represents the effect estimates of SNP on TGMV. The slope of the regression line is an estimate of the causal effect of BMI on TGMV. Based on SNP genetic instruments, a consistent causal relationship between higher BMI with lower TGMV was obtained using the IVW method (*β* = -1.53, se = 0.25, *P* = 8.20E-10), the weighted median method (*β* = -1.74, se = 0.36, *P* = 1.32E-06) and the MR–Egger regression (*β* = -1.86, se = 0.71, *P* = 9.39E-03). **B**. The horizontal axis represents different thresholds, = 0,0.05, 0.1,…, 0.5. The vertical axis represents the − value of the significance of the association analyses. valid-PRS_BMI_ maintained consistently significant negative associations with TGMV across all 10 thresholds (R < -0.047, P < 1.78E-07), valid-PRS_TGMV_ lost the significance with BMI in 10 thresholds (R > -0.016, P >0.083).

Further, using the newly developed MR-PRS approach [10], we obtained a consistent causal inference by eliminating the influence of pleiotropy. Specifically, in the independent second wave of neuroimaging data (*N* = 12867), we calculated the PRSs of BMI and TGMV (using the P-value threshold of 0.05), and both were significantly associated with the corresponding phenotypes (PRS_BMI_ with BMI: *R* = 0.232, *P* = 3.05E-149; PRS_TGMV_ with TGMV: *R* = 0.100, *P* = 6.33E-29). Further, we calculated the valid-PRSs using non-pleiotropic SNPs (see Materials and methods for details) and found correlations between these valid-PRSs and the corresponding main phenotypes remained significant (valid-PRS_BMI_ with BMI: R > 0.180, P < 6.24E-90; valid-PRS_TGMV_ with TGMV: R > 0.040, P < 7.79E-06), hence as qualified instrumental variables of randomized experiments. However, while valid-PRS_BMI_ demonstrated significant negative associations with TGMV consistently across all RT 10 thresholds (**Figure 2B**, R < -0.047, P < 1.78E-07), valid-PRS_TGMV_ was not associated with BMI at any of the 10 thresholds (**Figure 2B**, R > -0.016, P >0.083). The above results were reproducible using the mean-PRS averaged across 10 PT thresholds (from 0.05 to 0.5, by 0.05) or the best-fit-PRS instead of the P-value = 0.05 threshold (Supplementary Figure 1-2). Thus, following the argument of Mendelian randomization, we could conclude that an increased BMI will cause reduced TGMV, but not the other way around.

### Brain age prediction and comparison

We further investigated whether an increased BMI contributes to accelerated brain aging. The predicted brain age could be used to characterize the age-related brain atrophy. The brain-age prediction model was trained in the normal-weight group using XGBoost [27], and with a nested five-fold cross-validation (CV) framework, the model showed a moderate prediction accuracy in the normal-weight group (**Figure 3A**, *R* = 0.725, 95% CI=0.717-0.733, RMSE = 5.167, MAE = 4.130). As we expected, the brain-age gap was anti-correlated with the chronological age (**Figure 3B**, *R* = −0.698). The corrected brain-age got a large improvement after adjusted for bias (see Materials and methods for details) (**Figure 3C**, *R* = 0.897, 95% CI = 0.894–0.900, RMSE = 3.694, and MAE = 2.973). The trained model were then used to predict brain ages in the overweight and obesity group, and again we observed a high correlation between the chronological and predicted brain ages, after bias correction (*R* = 0.896, CI = 0.893–0.899, RMSE = 3.693, and MAE = 2.972; Supplementary Figure 3). We then compared the brain age gap (BAG) between the overweight/obesity group and the normal weight group. We found that the overweight group had a bigger BAG than the normal weight group with an average age increase of 0.590 years, *P* < 0.001, CI = 0.505-0.676, Cohen’s d = 0.139, and such a difference was even larger in the comparison between the obesity group and the normal-weight group, with an average age increase of 1.709 years (*P* < 0.001, CI = 1.602-1.815, Cohen’s d = 0.440).

**Figure 3.**
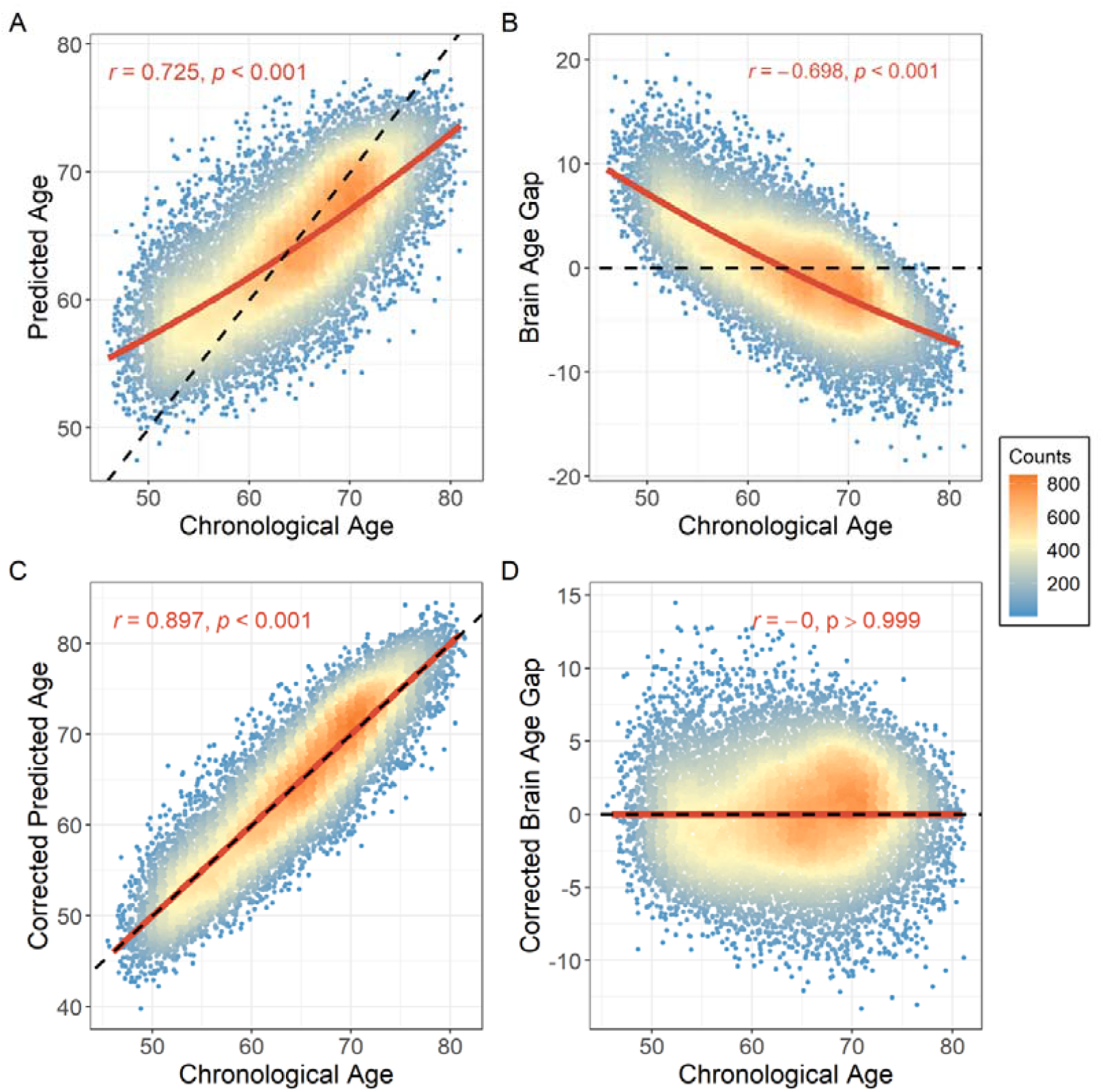
Performance of the brain-age prediction model in the normal-weight group. **A**. the predicted brain age was positively correlated with the chronological age (*R* = 0.725). **B**. The BAG was inversely correlated with the chronological age (*R* = −0.698). **C**. After bias adjusted, the corrected brain-age was correlated more with chronological age (R = 0.897). **D**. the corrected brain-age gap was orthogonal to the chronological age (R ≈ 0). The slope of the black dashed line was set to 1 in A and C to compare the prediction accuracy of the model. The slope of the black dashed line was set to 0 to show the deviation of brain age. The solid red line is the linear and quadratic fits to the chronological age in A and B, and the linear fit to the chronological age in C and D.

In order to further test the stability of this prediction pipeline, we retrained the prediction model with a 5-fold CV using all individuals (i.e. combining normal-weight and overweight/obesity groups), and found that the predicted brain age using the two pipelines were highly correlated (*R* = 0.985), indicating that our prediction results were highly stable.

### Association between BMI, BAG and cognitive function

We further explore whether BMI, BMI-related brain atrophy and TGMV have any implications on cognitive function. We first observed a negative correlation between BMI and cognitive function (**Figure 4A**, *R* = -0.072, *P* = 9.65E-26). We also found that both TGMV and the BAG were anti-correlated with cognitive function (*R* = -0.083, *P* < 0.001; *R* = -0.093, *P* = 3.93E-42, **Figure 4B**), while there was a positive correlation between BAG and the BMI (**Figure 4C**, *R* = 0.174, *P* = 1.1E-144). We further performed mediation analyses to assess whether TGMV and BAG contributed to the association between BMI and cognitive function. We conducted three mediation pathway analyses, namely 1) BMI → TGMV → BAG → cognitive function, 2) BMI → TGMV → cognitive function, 3) BMI → BAG → cognitive function. The results of the first model demonstrated that the indirect pathway of the association between BMI and cognitive function via TGMV and BAG was significant (**Figure 4D**, *β* = -0.013, *P* < 1E-04, and proportion of mediation: 18.06%). The results of the second model suggested that TGMV significantly mediated the association between BMI and cognitive function, that is, a moderate proportion of the relationship could be explained by TGMV (**Figure 4D**, *β* = -0.020, *P* < 1E-04, and proportion of mediation: 27.78%). The results of the third model showed that BAG significantly mediated the association between BMI and cognitive function (**Figure 4D**, *β* = -0.014, *P* < 1E-04, and proportion of mediation: 19.44%).

**Figure 4.**
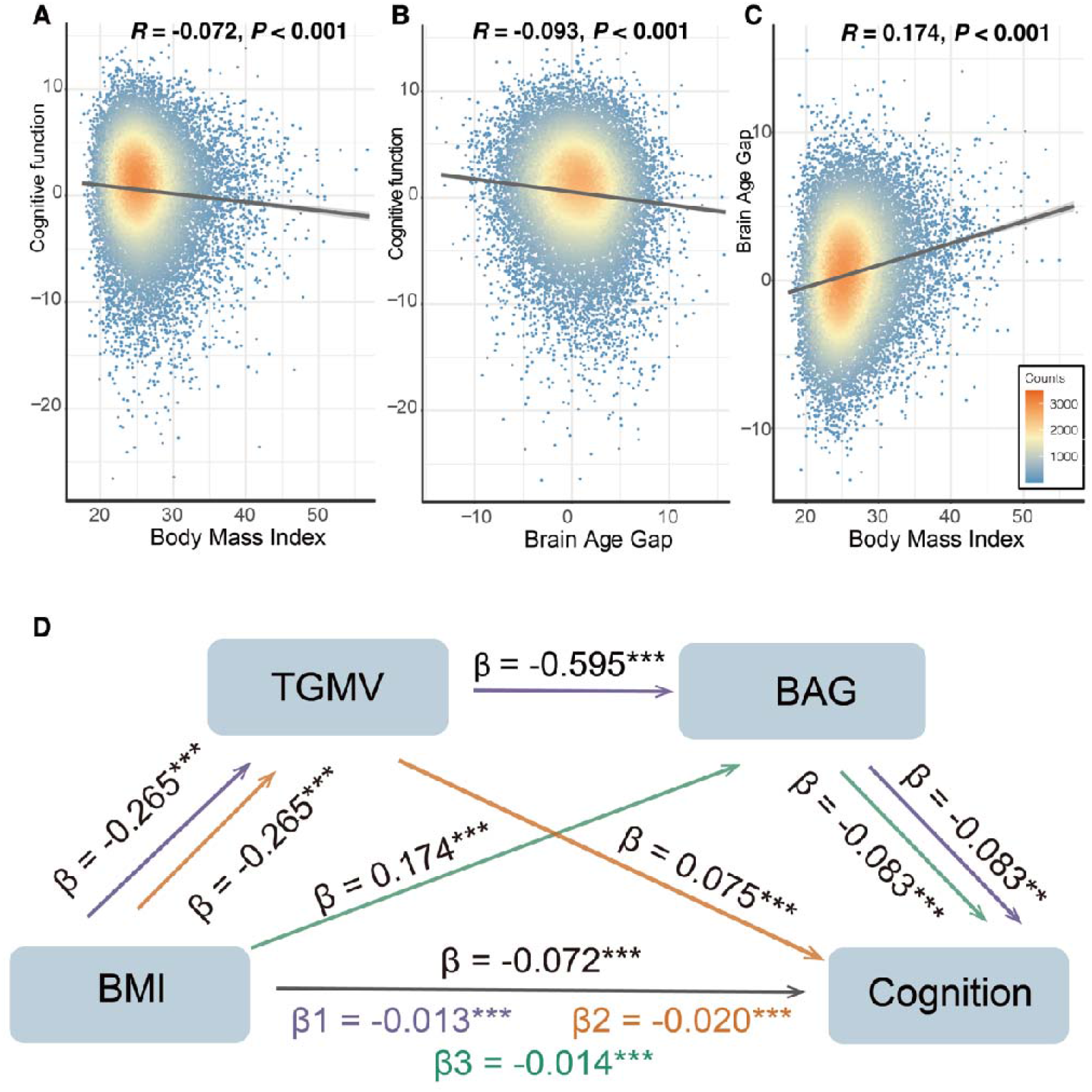
Results of mediation analyses. **A**. BMI was negative correlated with cognition function (*R* = -0.072, *P* = 9.65E-26). **B**. BAG was inversely correlated with cognitive function (*R* = -0.093, *P* = 3.93E-42), **C**. BAG was positively correlated with BMI (*R* = 0.174, *P* = 1.1E-144). **D**. The indirect pathway of the association between BMI and cognitive function via TGMV and BAG was significant (*β* = -0.013, *P* < 1E-04, and proportion of mediation: 18.06%). TGMV significantly mediated the association between BMI and cognitive function (*β* = -0.020, *P* < 1E-04, and proportion of mediation: 27.78%). BAG significantly mediated the association between BMI and cognitive function (*β* = -0.014, *P* < 1E-04, and proportion of mediation: 19.44%).

### Transcriptional Associates of BMI-linked structural association map

Finally, a remaining question of interest lies in the potential neurobiological mechanism underlying the association between BMI and GMV, and an intuitive approach is to investigate whether genetics play a role. We first performed a GWAS for BMI to identify BMI-associated genes. We identified 848 significant SNPs representing 295 independent loci, which were further mapped to 1342 genes based on position and cis-eQTL mapping (see Supplementary Methods for detail). To test whether BMI-associated genes may affect the brain, we narrowed these genes down to 380 based on cis-eQTL in the brain. We then examined the correlations between the spatial expression of the 314 BMI genes in the brain (314 out of 380 genes were available in the Allen Institute for Brain Science [21]) and the BMI-GMV association map. We hence identified eight validated significant genes (P_permute_ < 0.05 in the discovery sample and P_permute_ < 0.05 in the validation sample), including the TRIM27 gene, which is one of the TRIM family proteins and regulates tumor necrosis factor-alpha-induced apoptosis [31] (Table 2). In addition, we found 1027 BMI-associated genes (i.e. not mapped by cis-eQTL in the brain, or it’s expression not correlated to the BMI-linked map), of which the spatial expression was not associated with the BMI-GMV association map, and these genes were enriched in pathways involved in autophagy (**Figure 5**).

**Table 1.**
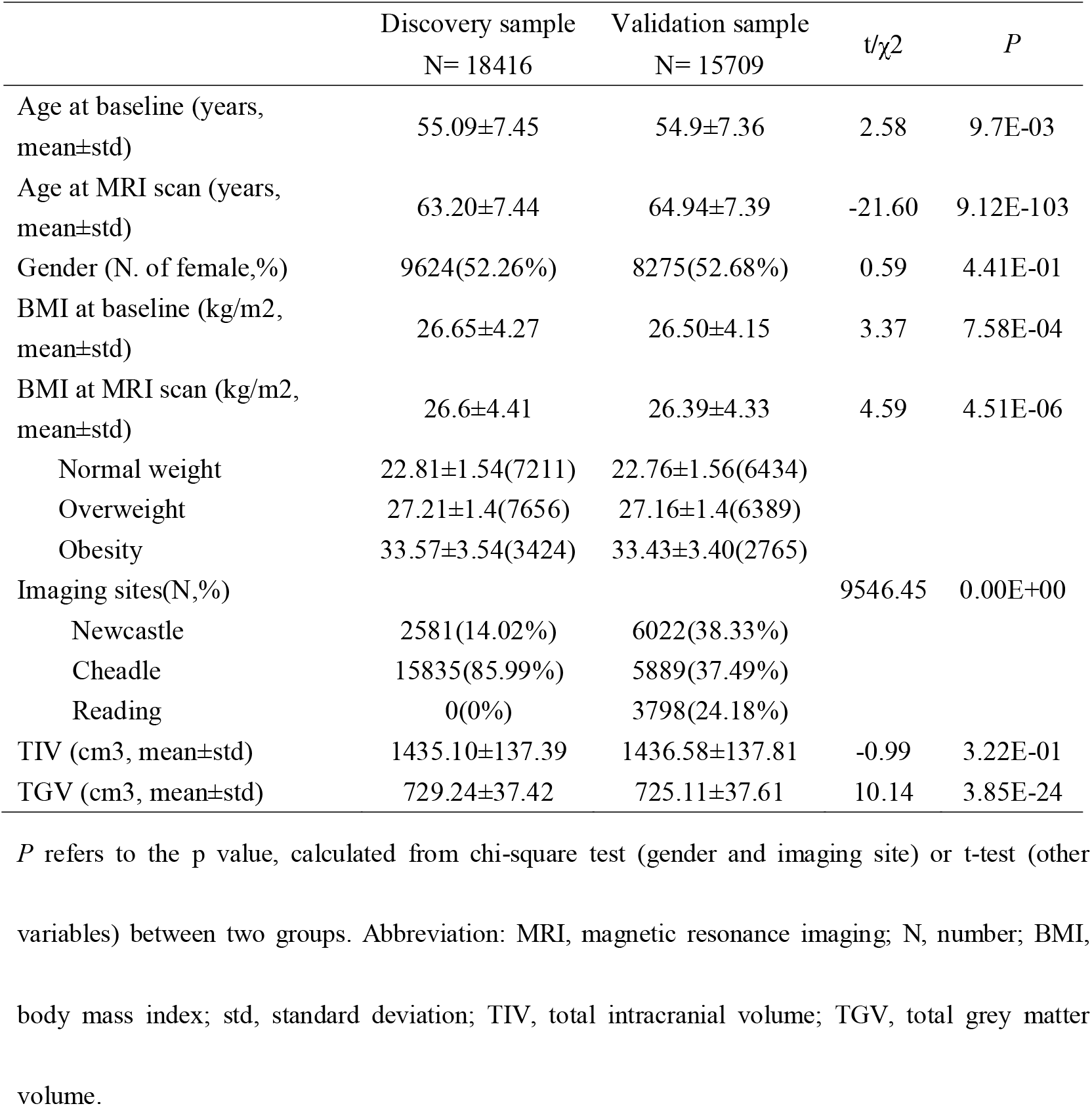
Demographics of study participants.

**Table 2.**
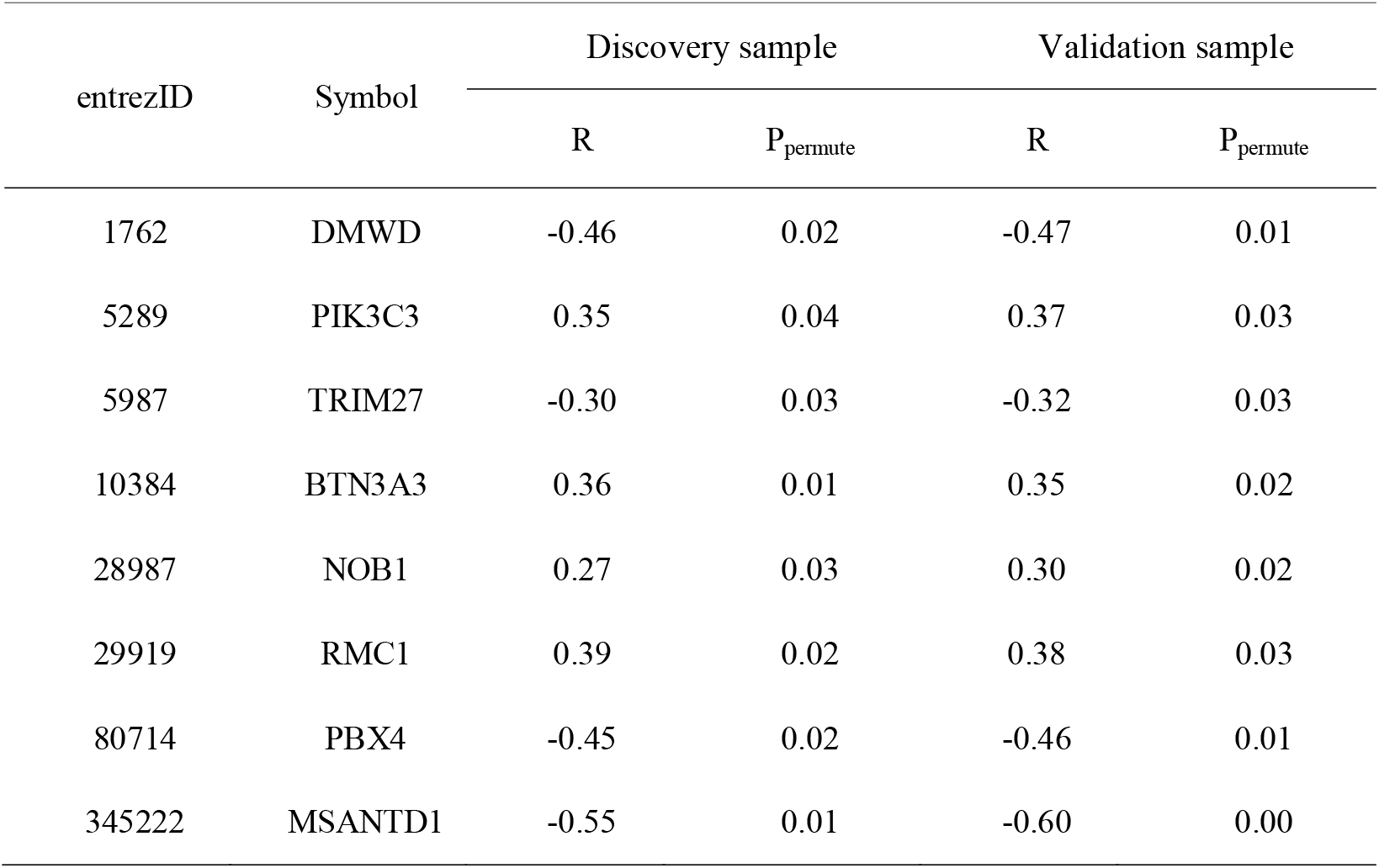
BMI-associated genes which were significantly associated with the BMI-linked structural association map.

| entrezID | Symbol | Discovery sample |  | Validation sample |  |
| --- | --- | --- | --- | --- | --- |
|  |  | R | P<sub>permute</sub> | R | P<sub>permute</sub> |
| 1762 | DMWD | -0.46 | 0.02 | -0.47 | 0.01 |
| 5289 | PIK3C3 | 0.35 | 0.04 | 0.37 | 0.03 |
| 5987 | TRIM27 | -0.30 | 0.03 | -0.32 | 0.03 |
| 10384 | BTN3A3 | 0.36 | 0.01 | 0.35 | 0.02 |
| 28987 | NOB1 | 0.27 | 0.03 | 0.30 | 0.02 |
| 29919 | RMC1 | 0.39 | 0.02 | 0.38 | 0.03 |
| 80714 | PBX4 | -0.45 | 0.02 | -0.46 | 0.01 |
| 345222 | MSANTD1 | -0.55 | 0.01 | -0.60 | 0.00 |

**Figure 5.**
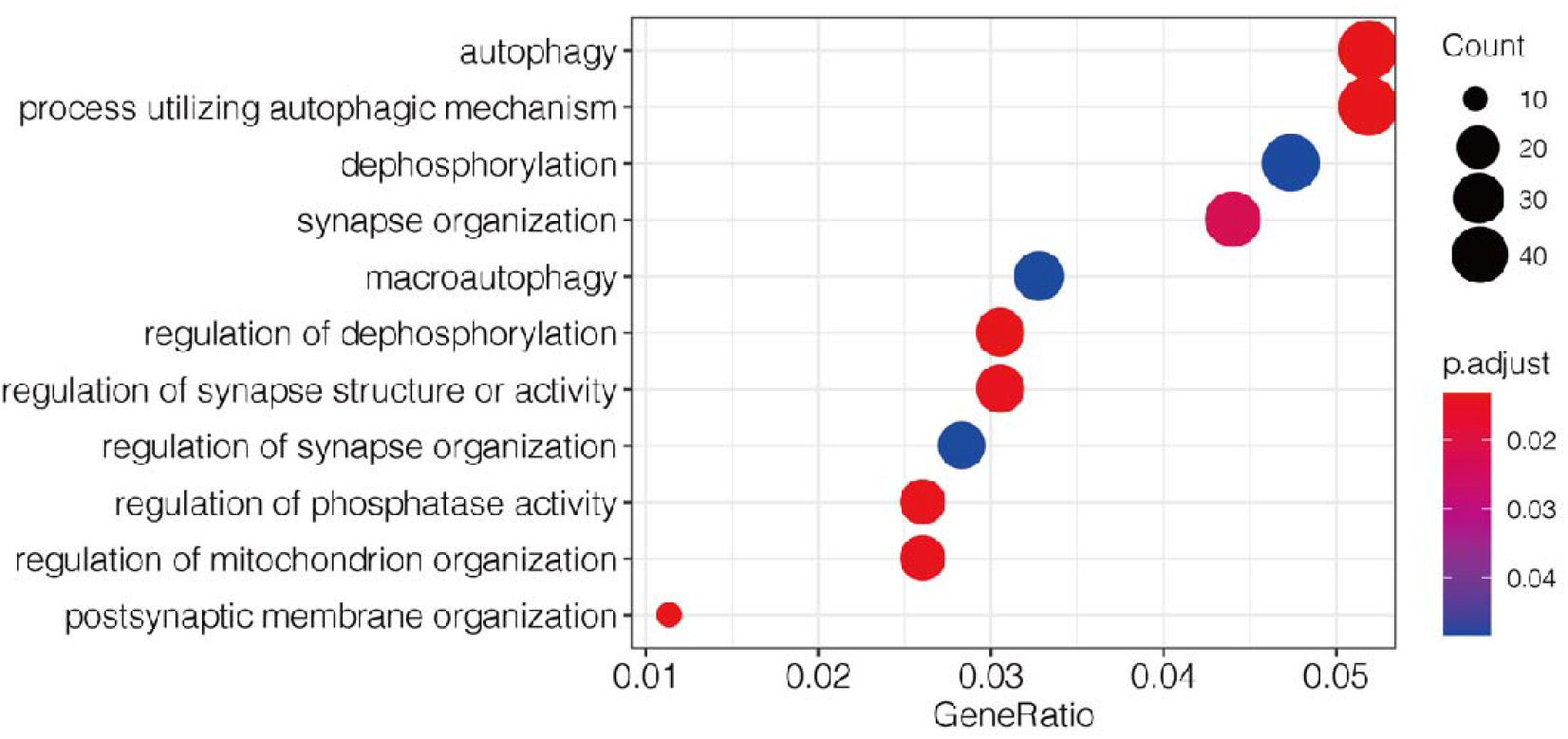
Function annotation of the BMI-associated genes. GO enrichment analyses of the 1027 BMI-associated genes, of which the spatial expression was not associated with the BMI-linked structural association map. Biological process GO terms were enriched.

## Discussion

Here, using multiple Mendelian randomization methods, we established and verified the potential harmful effects of higher body mass indices (BMIs) on brain grey matter volume in a large sample from the UK Biobank. Further, obese individuals exhibited accelerated brain aging and had a larger brain age gap (BAG) of 1.7 years. The brain age gap and TGMV explained 18% of the association between higher BMI and poorer cognitive function. In addition, pleiotropy of genes, e.g. TRIM27, may contribute to the relationship between BMI and brain atrophy. These results could improve the understanding of molecular mechanisms underlying obesity and related brain atrophy.

The pace of brain aging varies among individuals. Studying inter-individual differences in brain aging can help identify age-related health risks and develop personalized treatments. Estimates of the “brain age gap” are increasingly being used as biomarkers of brain health and brain degeneration [15-19]. Here, using a resource-efficient model XGBoost [27], in combination with a correction [32] to provide an unbiased brain age predictor, we for the first time explored the effects of obesity on brain aging, and found that obesity could accelerate age-related brain shrinkage. While individuals with excess weights had an averagely accelerated brain age of 0.59 years (95% CI: 0.505-0.676), the obese individuals had a larger acceleration of 1.7 years (95% CI: 1.602-1.815). However, the magnitude of accelerated brain age associated with obesity was found smaller in patients with neurological diseases such as Alzheimer’s disease [33] (5.76 years) and traumatic brain injury than in healthy controls [34] (4.66 years). Therefore, explanations for the above findings should not be exaggerated.

Obesity has been previously associated with brain-volume reduction [5,6]. Here, our study used multiple Mendelian randomization methods and showed evidence of unidirectional causality of BMI over brain volume, but not the other way around. However, the exact biological mechanisms by which obesity is associated with brain atrophy are complex and remain unclear. In our study, genes involved in biological functional pathways, including autophagy, were significantly enriched in BMI genes, of which the spatial expression pattern was not related to the BMI-GMV association map. Autophagy is a ubiquitous cellular process regulating cell growth, survival, development and death [35]. Previous literature has demonstrated that loss of autophagic homeostasis in adipose tissue has detrimental effects on metabolism and promotes metabolic disorders, while obesity may, in turn, lead to dysregulated autophagy [36]. Further, it has also been reviewed that autophagy inhibition may contribute to brain aging [37]. These results suggest that autophagy may be a biological mechanism underlying the causal relationship between obesity and brain atrophy.

In addition to the causal inference, our results further provide a complementary pleiotropy explanation for the relationship between obesity and brain atrophy. Spatial gene expression of the TRIM27 gene negatively correlated with the BMI-linked GMV reduction throughout the brain, implying that the TRIM27 gene is mostly expressed in brain regions associated with BMI. TRIM27 gene encodes a member of the tripartite motif (TRIM) family involved in many biological processes, including cell differentiation and apoptosis signaling [38]. Previous studies demonstrated that induction of adipocyte apoptosis removes adipocytes [39]. A study reported that, in a neurotoxin model of Parkinson’s disease, TRIM27 deficiency reduces apoptosis of dopaminergic neurons [40]. TRIM27 knockdown significantly inhibits Glu-mediated apoptosis [41]. Our results indicated that some genetic variants might increase the risk for obesity and simultaneously promote brain volume reduction through promoting apoptosis. These lines of evidence suggest that some pleiotropic genetic variants may alter obesity risk and simultaneously affect the expression of genes in the brain, leading to brain atrophy.

Our findings have to be considered in the context of the following limitations. First, this study is based on the elderly aged 45-81 years, and previous literature has reported an increased obesity rate in elderly individuals [42], and hence our results still need to be replicated in younger individuals. Second, our study’s brain age prediction model solely used structural MRI data, and combined with other modalities (e.g., diffusion MRI and functional MRI), it may achieve higher accuracy. Nevertheless, structural MRI may capture more aging information than other MRI modalities [43]. Finally, the detailed neurobiological mechanisms of how pleiotropic genes function in the brain remain unclear and require further study.

In conclusion, an increased BMI has pernicious effects on the brain’s gray matter volume (GMV). Genes involved in autophagy and genes, including TRIM27, may contribute to the negative association between BMI and brain GMV. This study provides a comprehensive understanding of the relationship between genetics, obesity, brain atrophy, aging, and cognition, which altogether can help to develop personalized treatments for adiposity to minimize the side effects of excess obesity on the brain and, maybe more importantly, provide a potential new research direction in revealing the neurobiological mechanisms behind the obesity-brain connection.

## Data Availability

All UK Biobank data used in this work were obtained under Data Access Application 19542 and are available to eligible researchers through the UK Biobank (www.biobank.ac.uk). Gene expression data from the Allen Institute for Brain Science are freely available at https://human.brain-map.org/static/download.

## Code Availability

Statistical Parametric Mapping package (SPM12) and the voxel-based morphometry 8 (VBM8) toolbox were used to process the imaging data. PLINK 1.90 and PRSice (www.PRSice.info) were used to perform genome-wide association analysis and calculate the polygenic risk score respectively. FUMA online platform (http://fuma.ctglab.nl/) was used to perform SNP-gene mapping. TwoSampleMR R package (https://github.com/MRCIEU/TwoSampleMR) was used to perform two-sample mendelian randomization analysis. R package lavaan version 0.8 was used to perform mediation analysis. R package XGBoost (https://github.com/dmlc/xgboost) was used to train a brain age prediction model.

## Funding

**J.F**. is supported by National Key R&D Program of China (No. 2018YFC1312904 and No. 2019YFA0709502), the Shanghai Municipal Science and Technology Major Project (No. 2018SHZDZX01), the 111 Project (No. B18015), Shanghai Center for Brain Science and Brain-Inspired Technology and Zhangjiang Lab.

**T.J**. is supported by National Key R&D Program of China (No. 2019YFA0709501) and the National Natural Science Foundation of China (T2122005, No. 81801773).

**S.G**. is supported by the National Natural Science Foundation of China (NSFC) grant (No. 12071124).

The funders had no role in study design, data collection and analysis, decision to publish or preparation of the manuscript.

## CRediT authorship contribution statement

**J.K**.: Conceptualization, Methodology, Formal analysis, Visualization, Writing - original draft, Writing -review & editing. **T.J**.: Supervision, Conceptualization, Methodology, Writing -review & editing, Funding acquisition. **L.Z**.: Formal analysis, Visualization. **Y.L**.: Formal analysis, Visualization. **W.C**.: Conceptualization, Formal analysis. **S.G**.: Conceptualization, Methodology, Formal analysis, Funding acquisition. **J.F**.: Supervision, Conceptualization, Writing -review & editing, Project administration, Funding acquisition.

## Acknowledgments

The authors would like to thank UK Biobank participants for their time and UK Biobank team members for collating the data.

## Declaration of competing interest

The authors declare no conflict of interest with respect to the research, authorship, and publication of this article.

## Notes

### Competing Interest Statement

The authors have declared no competing interest.

### Author Declarations

The UK Biobank study received ethical approval from the NHS National Research Ethics Service North West (reference number: 16/NW/0274). Data access permission was granted under UKB application 19542. All necessary patient/participant consent has been obtained and the appropriate institutional forms have been archived.

