## supplementary materials for "Association between obesity, brain atrophy and accelerated brain aging and their genetic mechanisms"

### Supplementary Materials and Methods

#### Cognitive function

UK Biobank assessed a range of cognitive functions (https://biobank.ctsu.ox.ac.uk/showcase/label.cgi

?id=100026) including fluid intelligence (Field 20016), numeric memory (Field 4282), prospective memory (Field 20018), reaction time (Field 20023), pairs matching (Field 399), matrix pattern completion (Field 6373), symbol digit substitution (Field 23324), tower rearranging (Field 21004) and trail making (Field 6348 and 6350). We used the sum of the normalized score of the above items. Higher scores indicated better cognitive performance.

#### Structural MRI data acquisition and preprocessing

Structural MRIs were collected across three imaging centres that were equipped with identical scanners (Siemens Skyra 3T). Structural images were acquired with straight sagittal orientation with a resolution of 1 × 1 × 1 *mm* and a field of view of 208 × 256 × 256 matrix, over a duration of 5 minutes, and with 1-mm isotropic resolution using a 3-dimensional magnetization-prepared rapid-acquisition gradient echo. The MRI protocols have been described in detail elsewhere [1].

The structural MRI data were preprocessed in the Statistical Parametric Mapping package (SPM12) [2] using the voxel-based morphometry 8 (VBM8) toolbox with default settings, including the usage of high-dimensional spatial normalisation with an already integrated Dartel template in Montreal Neurological Institute (MNI) space. All images were subjected to nonlinear modulations and corrected for each individual head size. Images were then smoothed with a 6 *mm* full-width at half-maximum Gaussian kernel with the resulting voxel size 1.5 *mm*^3^. The estimated total intracranial volume (TIV) was calculated as the summation of the grey matter, white matter, and CSF volume in native space. The automated anatomical labelling 3 (AAL3) atlas [3], which partitioned the brain into 166 regions of interest, was employed to obtain the total brain grey matter volume and region-wise grey matter volume.

#### Genetic data quality control

Genotyped data are available for all 500,000 participants in the UKB cohort. All blood samples were genotyped using the UK BiLEVE array and the UK Biobank axiom array, which have over 95% common marker content. Details of the array design, genotyping, quality control and imputation are available in a previous publication [4].

In this study, we performed stringent quality control standards using PLINK 1.90 [5]. Specifically, single-nucleotide polymorphisms (SNPs) with call rates < 95%, minor allele frequency < 0.1%, deviation from the Hardy–Weinberg equilibrium with p < 1E-10 were excluded from the analysis. In addition, perform sample quality control based on missingness, relatedness, gender mismatch, non-British ancestry using the sample quality control information provided by UK Biobank. Finally, we have 336495 samples with 667199 variants.

#### Preprocessing of the Allen Human Brain Atlas data

We followed the preprocessing pipeline suggested by Arnatkevic̆iūtė et al. [6] and using the same pipeline as previous studies [7], including probe-to-gene re-annotation, data filtering, probe selection. We separated the samples into the areas defined by the AAL3 atlas based on their MNI coordinates and excluded the samples located outside of the grey matter defined by this atlas. To control for the inter-individual differences, we conducted within-sample and across-gene normalisation. One gene failed the normalisation and therefore was deleted, resulting in 15,408 genes. The mean across gene expression of all the samples located in the brain region and the mean of gene expression of all the subjects were used as the mean expression of each gene in each brain region.

#### Genome-wide association analysis and variants annotation

We performed genome-wide association analysis (GWAS) of BMI using PLINK 1.90 [5], with covariates including age, gender and the top 40 genetic principal components. We used the FUMA [8] software to map the independent significant SNPs to genes based on positional and eQTL mapping. Positional mapping map SNPs to genes based on physical distance (within a 10-kb window). The eQTL mapping map SNPs to genes based on eQTL associations that SNP was significant (false discovery rate (FDR)≤ 0.05) expression level of gene using information on eQTLs of 49 tissue type in GTEX [9] v8. The eQTL mapping was based on cis-eQTLs and could map SNPs to genes up to 1Mb apart. We also used the cis-eQTL information from GTEX [9] v8 and BrainNEAC [10] databases to ascertain genes of which the expression in the brain was associated with SNP.

### Supplementary Figures

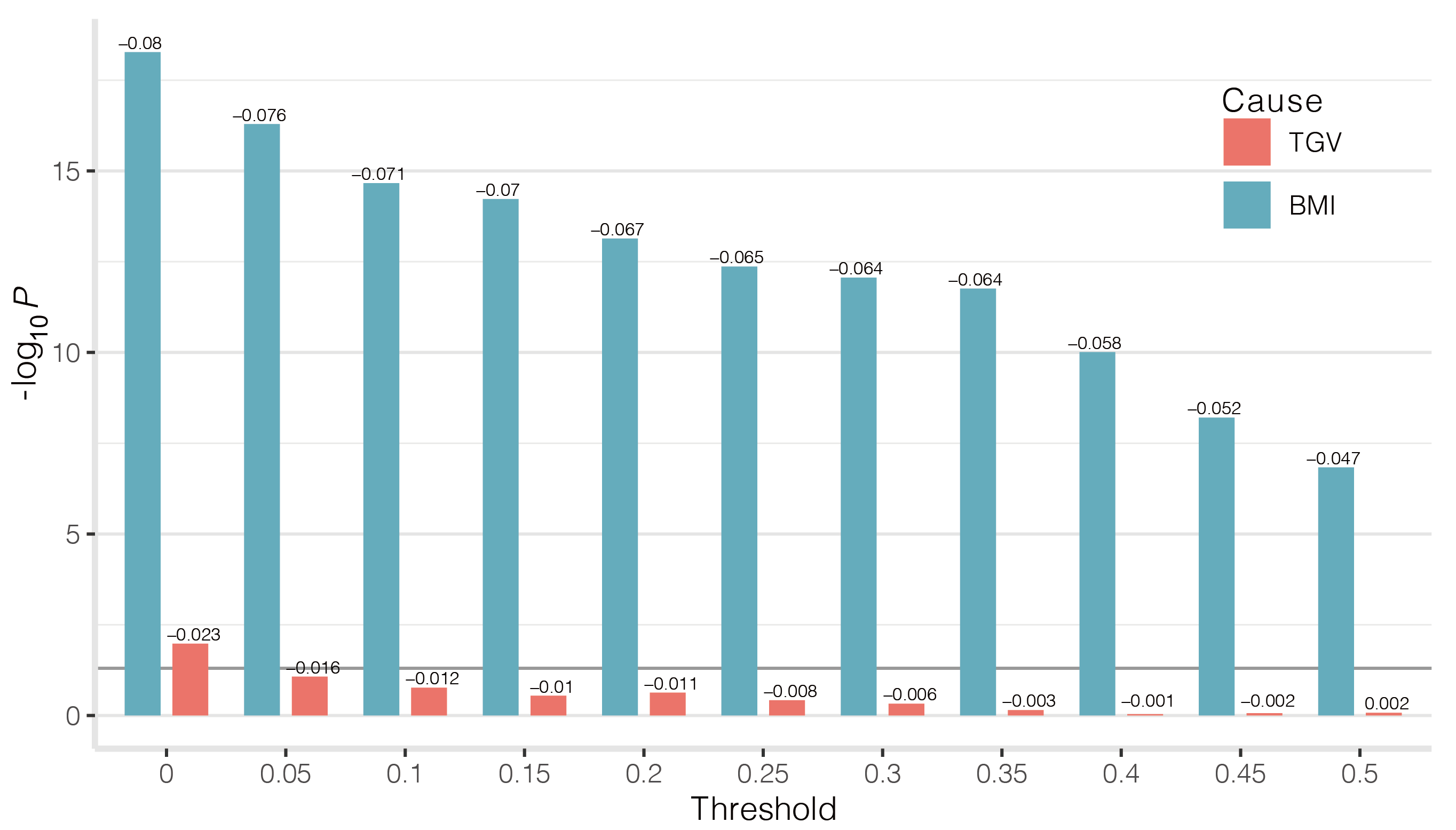

**Supplementary Figure 1. Results of MR-PRS method when using the mean-PRS averaged across 10 thresholds (from 0.05 to 0.5, by 0.05).** The horizontal axis represents different 𝑅𝑇 thresholds, 𝑅𝑇 = 0,0.05, 0.1,..., 0.5. The vertical axis represents the −𝑙𝑜𝑔 𝑃 value of the significance of the association analyses.

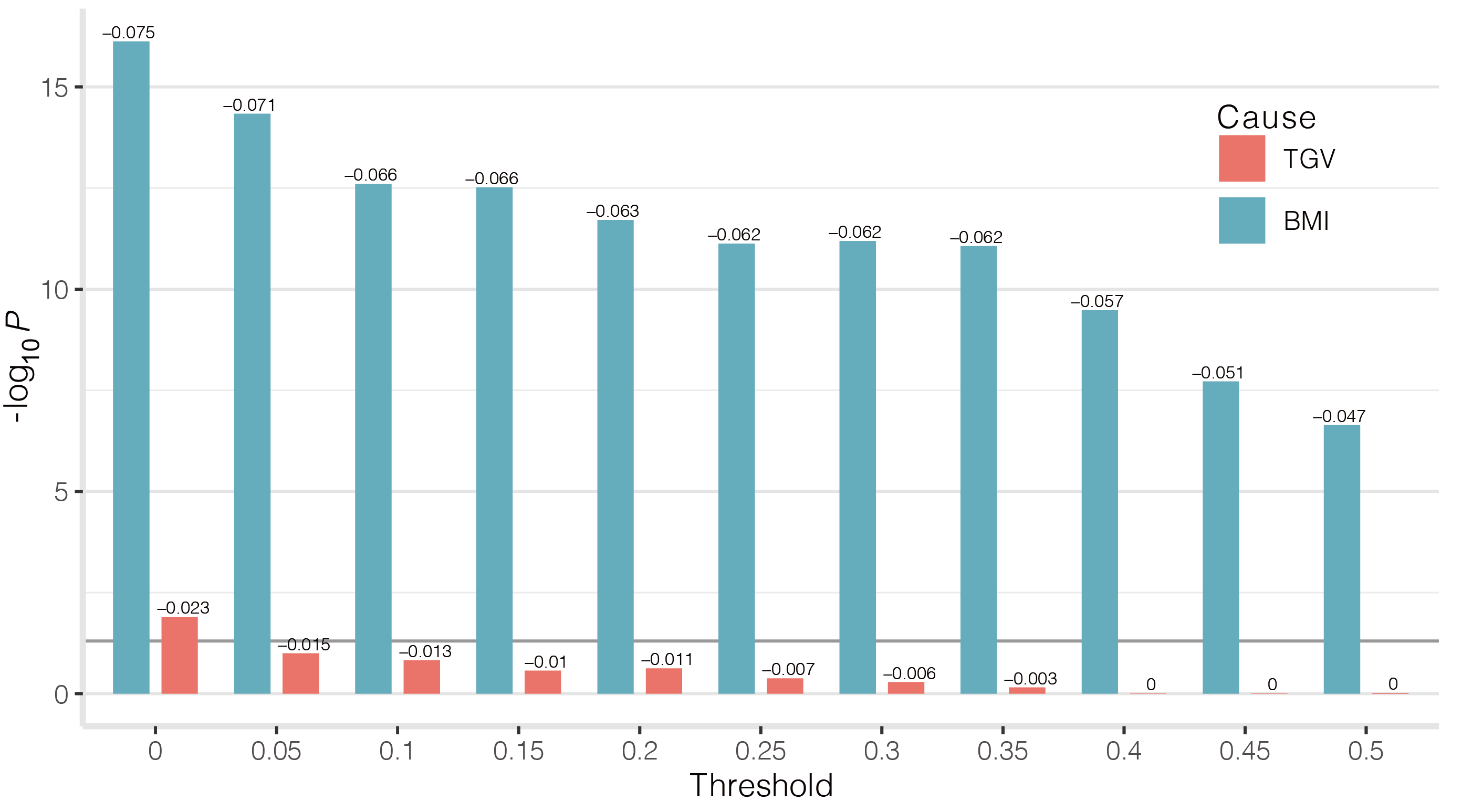

**Supplementary Figure 2. Results of MR-PRS method when using the best-fit-PRS across 10 thresholds (from 0.05 to 0.5, by 0.05).** The horizontal axis represents different 𝑅𝑇 thresholds, 𝑅𝑇 = 0,0.05, 0.1,..., 0.5. The vertical axis represents the −𝑙𝑜𝑔 𝑃 value of the significance of the association analyses.

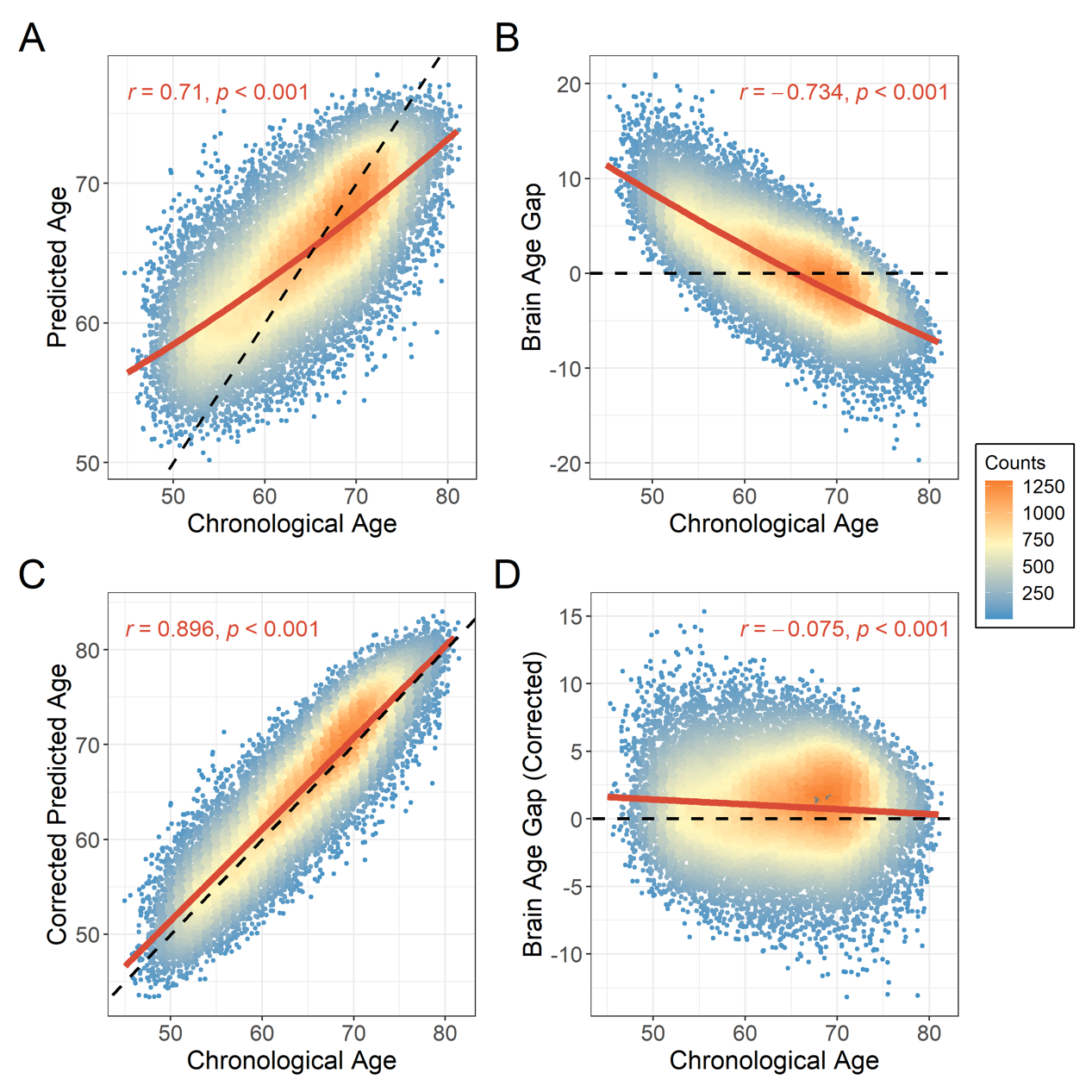

**Supplementary Figure 3. Performance of the brain-age prediction model in the over-weight and obese group.** **A.** the predicted brain age was positively correlated with the chronological age (*R* = 0.71). **B.** The BAG was inversely correlated with the chronological age (*R* = −0.734). **C.** After bias adjusted, the corrected brain-age was correlated more with chronological age (R = 0.896). **D.** the corrected brain-age gap was orthogonal to the chronological age (R = -0.075). The slope of the black dashed line was set to 1 in A and C to compare the prediction accuracy of the model. The slope of the black dashed line was set to 0 to show the deviation of brain age. The solid red line is the linear and quadratic fits to the chronological age in A and B, and the linear fit to the chronological age in C and D.
